# Analgesic Equivalence of NSAIDs and a Weak Opioid in Acute Postoperative Pain: A Randomised Controlled Trial

**DOI:** 10.64898/2026.05.03.26352343

**Authors:** Paola E. Vallejo-Mora, Phabel A. Lopez-Delgado, Mirna M. Delgado-Carlo

## Abstract

**Background:** Non-steroidal anti-inflammatory drugs (NSAIDs) and weak opioids such as tramadol are cornerstones of multimodal analgesia, particularly in settings with limited access to potent opioids. However, cross-class equianalgesic data are scarce. This randomized controlled trial evaluated the analgesic equivalence of tramadol, ketorolac, and diclofenac as premedication in patients undergoing minimally invasive surgery.

**Methods:** In this double-blind, parallel-group trial, 30 patients undergoing elective minimally invasive surgery under balanced general anaesthesia were randomised to intravenous tramadol 150 mg, ketorolac 60 mg, or diclofenac 150 mg 45 minutes before skin incision. The sample size was calculated using a one-way ANOVA to detect a large effect size (Cohen’s *f* = 0.4) between the three groups in the primary outcome (NRS pain score at 60 minutes), with *α* = 0.05 and 80% power, yielding 10 patients per group (30 patients total). Pain intensity was assessed using the Numeric Rating Scale (NRS, 0–10) and Verbal Rating Scale (VRS) at recovery room arrival (T0) and at 30 (T1), 60 (T2), and 90 (T3) minutes thereafter. Secondary outcomes included rescue morphine consumption and safety. Between-group comparisons used Kruskal–Wallis tests.

**Results:** All three groups converged to a median NRS of 2 by 60 minutes (T2), with no statistically significant differences in pain scores or rescue morphine requirements at T2 or T3 (p > 0.05). At T0 and T1, NRS scores were higher in the ketorolac group (median 1.5 and 3) compared with tramadol and diclofenac (both median 0 at T0; T1: tramadol 1, diclofenac 2; p < 0.05). Rescue morphine was required in 0/10 (tramadol), 3/10 (ketorolac), and 2/10 (diclofenac) (p = 0.19). No hypersensitivity reactions occurred.

**Conclusions:** Equianalgesic doses of tramadol, ketorolac, and diclofenac provided comparable postoperative pain control over 90 minutes following minimally invasive surgery. All three agents were well tolerated. These findings provide clinically useful guidance for analgesic selection in resource-limited settings.

**Trial registration:** ClinicalTrials.gov - NCT07500454 (retrospectively registered).

**Highlights:**

1. RCT (n=30) comparing equianalgesic doses of tramadol, ketorolac, and diclofenac in minimally invasive surgery.
2. All three groups converged to median NRS 2 by 60 minutes postoperatively.
3. Early higher pain in ketorolac group partly attributed to age imbalance (*ρ* = 0.49, *p* = 0.006).
4. No hypersensitivity reactions occurred; all agents were well tolerated.
5. These findings support the use of any of these agents as premedication in minimally invasive surgery.

**Figure 1: Graphical Abstract – CONSORT**

Figure 1:
Graphical Abstract.
CONSORT flow diagram showing participant recruitment, randomization (n=30), and follow-up (100% completion). Study design comparing three premedication regimens: tramadol 150 mg, ketorolac 60 mg, and diclofenac 150 mg administered intravenously 45 minutes before skin incision. Key finding: all three groups converged to a median NRS 2 by 60-90 minutes postoperatively. Made with *BioRender*.

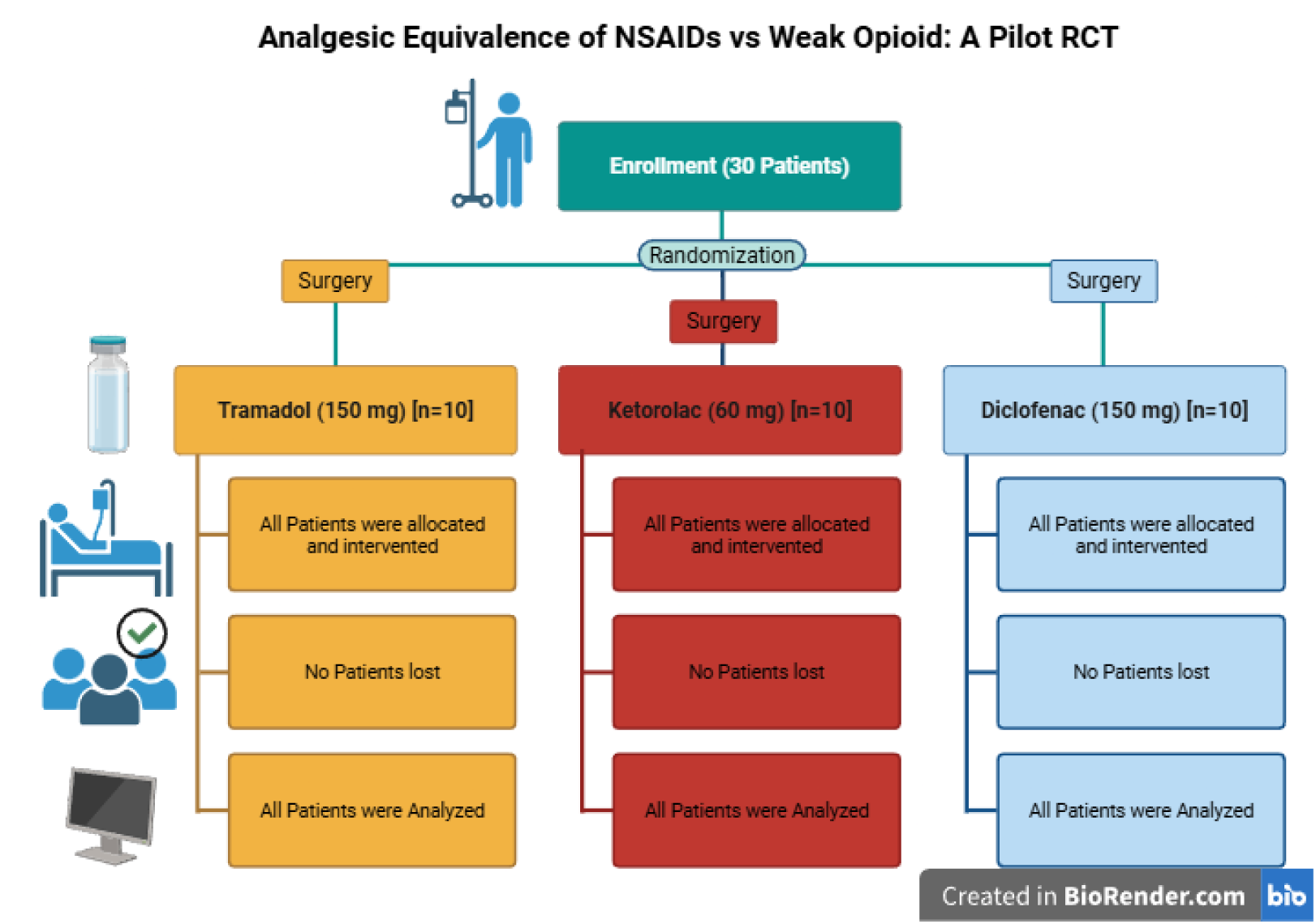

## 1. Introduction

Acute postoperative pain is among the most frequent and burdensome complications of surgical care. More than half of patients who experience postoperative pain report that it was inadequately treated, with persistent pain driving increased perioperative morbidity and mortality, prolonged hospital stay, higher healthcare costs, and diminished quality of life [1, 2]. Sub-optimal analgesic dosing — whether through under- or over-administration — is a major contributing factor. For the practising anaesthesiologist, a thorough understanding of equianalgesic relationships between drug classes is therefore a fundamental clinical tool [3].

Analgesic equivalence between opioid agents has been well characterised and is widely applied in perioperative practice. By contrast, cross-class equianalgesic data comparing non-steroidal anti-inflammatory drugs (NSAIDs) with weak opioids remain limited. This gap is clinically relevant, as NSAIDs and weak opioids such as tramadol represent the backbone of multimodal analgesia protocols in many low- and middle-income settings where access to potent opioids is restricted [4, 5].

We conducted a randomized controlled trial to evaluate the analgesic equivalence of three commonly used perioperative analgesics: ketorolac (60 mg IV), diclofenac (150 mg IV), and tramadol (150 mg IV), administered at equianalgesic doses as premedication in patients undergoing elective minimally invasive surgery under balanced general anaesthesia. These doses were selected on the basis of established morphine-equivalence ratios (10 mg ketorolac *≈* 4 mg morphine *≈* 40 mg tramadol), extrapolated to single-dose premedication. The sample size was calculated using a one-way ANOVA to detect a large effect size (Cohen’s *f* = 0.4) between the three groups in the primary outcome (NRS pain score at 60 minutes), with *α* = 0.05 and 80% power, yielding 10 patients per group (30 patients total). We hypothesised that the three agents would provide comparable analgesia at equianalgesic doses. The study was conducted at the Hospital Regional “General Ignacio Zaragoza,” ISSSTE, Mexico City, and registered with the institutional research registry (RPI #403-2024).

## 2. Methods

### 2.1. Study design and ethics

This was a single-centre, prospective, double-blind, parallel-group randomized controlled trial. The study was approved by the Hospital’s Ethics and Research Committee and conducted in accordance with the Declaration of Helsinki and Mexican national research regulations (*Ley General de Salud*, Título Segundo). Written informed consent was obtained from all participants prior to enrolment.

### 2.2. Sample Size Calculation

The sample size was calculated using a one-way ANOVA to detect a large effect size (Cohen’s *f* = 0.4) between the three groups in the primary outcome (NRS pain score at 60 minutes), with *α* = 0.05 and 80% power. This yielded 10 patients per group (30 patients total).

### 2.3. Participants

Eligible patients were aged 18–55 years, ASA physical status I or II, with a body mass index between 18.5 and 34.99 kg/m^2^, scheduled for elective minimally invasive surgery under balanced general anaesthesia; specifically, 28 laparoscopic cholecystectomies and 2 laparoscopic abdominal wall repairs. Patients were excluded for pregnancy, known hypersensitivity to any study drug, pre-existing acute pain with NRS *≥* 4 or VRS *≥* “moderate” before surgery, or chronic pain with current analgesic use. Elimination criteria included withdrawal of consent before premedication, surgical duration exceeding 180 minutes, conversion from laparoscopic to open approach, any hypersensitivity reaction during drug administration, haemodynamic shock of any aetiology, or requirement for postoperative mechanical ventilation due to anaesthetic-surgical complications.

### 2.4. Randomization and blinding

Thirty patients were randomised equally to three groups (*n* = 10 each) using simple random allocation with numbered assignments. Study drugs were prepared in identical 100 mL 0.9% saline bags by personnel not otherwise involved in the study and delivered unlabelled to the responsible anaesthesiologist, ensuring double blinding of both investigator and patient.

### 2.5. Interventions

All patients received balanced general anaesthesia: induction with fentanyl 4 mcg/kg, vecuronium 100 mcg/kg, and propofol 1.5 mg/kg; maintenance with fentanyl infusion at 1–2.5 mcg/kg/h. Paracetamol, additional NSAIDs, lidocaine infusions, and magnesium sulphate infusions were not used during the procedure. Study drugs were administered intravenously 45 minutes before skin incision: Group TRAM received tramadol 150 mg IV, Group KETO received ketorolac 60 mg IV, and Group DICLO received diclofenac 150 mg IV, each as a single daily dose.

### 2.6. Outcomes and measurements

The primary outcome was pain intensity, assessed using the Numeric Rating Scale (NRS, 0–10) at 60 minutes post-recovery arrival (T2). The NRS and the Verbal Rating Scale (VRS: 0 = absence; 1 = low; 2 = moderate; 3 = severe) [6] were measured at five time points: pre-surgical baseline (Pre), recovery room arrival (T0), and at 30 (T1), 60 (T2), and 90 (T3) minutes after T0.

Secondary outcomes included:

- Rescue analgesia: number of patients requiring morphine 3 mg IV (on patient request or clinical indication) and total morphine consumption (mg) at each interval.
- Safety: hypersensitivity reactions classified according to Müller criteria and managed per institutional protocol (hydrocortisone 100 mg IV; adrenaline 0.5 mg IM for anaphylaxis) [7].

### 2.7. Statistical analysis

Descriptive statistics are reported as median (range) for continuous variables and as frequencies with percentages for categorical variables. Between-group comparisons of NRS scores were performed using the Kruskal–Wallis test with Dunn post-hoc tests and Holm correction for multiple comparisons; effect size was estimated as ordinal epsilon-squared (*ε*^2^). Within-group NRS trajectories were analysed using the Friedman test with Durbin–Conover post-hoc testing and Holm correction; concordance was quantified by Kendall’s *W* with 95% confidence intervals. VRS and rescue analgesia data were analysed using Pearson’s chi-squared test with Cramér’s *V* as effect size.

All analyses were performed in R statistical software (version 4.3.1 (2023-06-16 ucrt)), using the *ggstatsplot* package [8]. A two-sided *p <* 0.05 was considered statistically significant.

### 2.8. Trial Registration

This trial was prospectively registered with the Instituto de Seguridad y Servicios Sociales de los Trabajadores del Estado (ISSSTE) under registration **ISSSTE RPI #403-2024**. The trial was retrospectively registered on ClinicalTrials.gov (ID: **NCT07500454**). All outcomes are reported in accordance with CONSORT guidelines.

## 3. Results

### 3.1. Recruitment and Retention

A total of 30 patients were enrolled over a 1-month recruitment period (7.5 patients per week). All 30 participants completed the 90-minute follow-up (retention rate 100%), and every patient received the allocated intervention without major protocol deviations (protocol adherence 100%).

### 3.2. Participant Characteristics

Thirty patients were enrolled and completed the study (10 per group); no participants were eliminated after randomization. Baseline characteristics are presented in Table 1. The overall sample had a median age of 29 years (range 19–55) and included patients of both sexes and ASA I–II status. Surgical duration was similar across groups (Group TRAM: median 110 min; Group KETO: 120.5 min; Group DICLO: 123.5 min). All patients presented with NRS = 0 and VRS = “absence” at the pre-surgical baseline (Pre), confirming homogeneous baseline pain status across groups. The ketorolac group had a numerically higher mean age (44.5 years) compared with the tramadol (32.2 years) and diclofenac (34.7 years) groups, which constituted an uncontrolled confounding variable (Table 1).

**Table 1:** Baseline characteristics by Group.

| Variable | Tramadol 150 mg<br>(*n* = 10) | Ketorolac 60 mg<br>(*n* = 10) | Diclofenac 150 mg<br>(*n* = 10) |
| --- | --- | --- | --- |
| Age, mean (range), yr | 32.2 (19–48) | 44.5 (28–55) | 34.7 (20–54) |
| Sex, M/F | 7/3 | 5/5 | 6/4 |
| ASA I/II | 5/5 | 4/6 | 6/4 |
| BMI category (Normal) | 10/10 (100%) | 10/10 (100%) | 10/10 (100%) |
| Surgical duration, median, min | 110 | 120.5 | 123.5 |
| Anaesthetic duration, median, min | 131 | 138.5 | 145.5 |
| Pre-surgical NRS (TB) | 0 (all) | 0 (all) | 0 (all) |
| Pre-surgical VRS (TB) | Absence (all) | Absence (all) | Absence (all) |

### 3.3. Primary Outcome: NRS Pain Scores

Between-group comparisons of NRS scores at each time point are summarised in Table 2 and Figure 2. At T0, a statistically significant difference was detected across groups (Kruskal–Wallis *χ*^2^(2) = 8.88, *p* = 0.012, *ε*^2^ = 0.31), driven by higher NRS scores in the ketorolac group (median 1.5) compared with tramadol and diclofenac (both median 0; Dunn post-hoc, both *p* = 0.025). While all three groups had identical NRS = 0 at baseline, the ketorolac group developed greater pain intensity during anaesthetic emergence, which may reflect the higher mean age in that cohort rather than a direct drug effect. A similar pattern persisted at T1 (Kruskal–Wallis *χ*^2^(2) = 6.52, *p* = 0.038, *ε*^2^ = 0.23), where ketorolac scores remained higher than tramadol (*p* = 0.033). By T2 and T3, all between-group differences resolved, with identical median NRS of 2 across all three groups (T2: *p* = 0.809; T3: *p* = 0.271).

**Figure 2:**
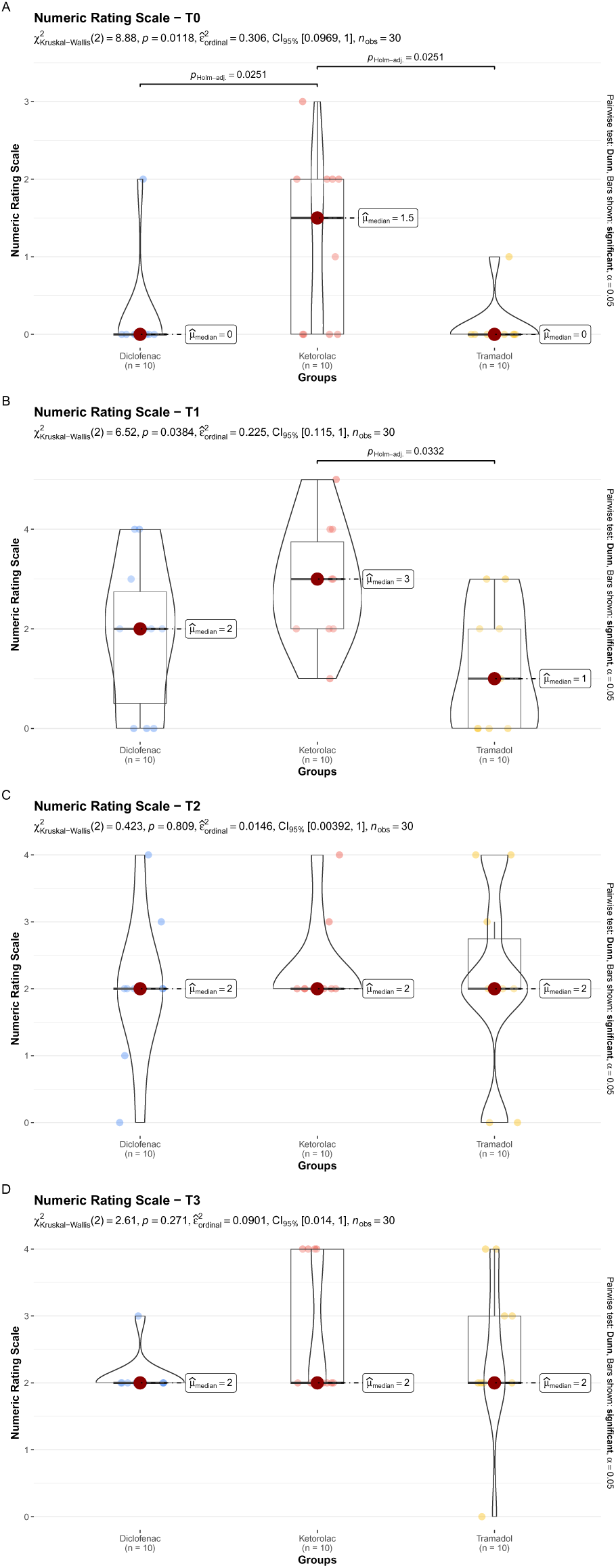
Pain Intensity (NRS) Over Time by Treatment Group. Box plots show median, interquartile range, and range for diclofenac (green), ketorolac (red), and tramadol (blue) at recovery room arrival (T0) and at 30 (T1), 60 (T2), and 90 (T3) minutes. All groups converged to median NRS 2 by T2–T3. *p < 0.05 for between-group comparisons (Kruskal–Wallis with Dunn post-hoc).

**Table 2:** NRS pain scores by group and time point (median, range).

| Time | Tramadol | Ketorolac | Diclofenac | KW<br>*p*-value | $\varepsilon^2$ | Post-hoc |
| --- | --- | --- | --- | --- | --- | --- |
| T0 | 0 (0–2) | 1.5 (0–4) | 0 (0–2) | 0.012 | 0.31 | KETO > TRAM, DICLO* |
| T1 (30 min) | 1 (0–3) | 3 (0–5) | 2 (0–4) | 0.038 | 0.23 | KETO > TRAM<sup>†</sup> |
| T2 (60 min) | 2 (0–4) | 2 (0–4) | 2 (0–4) | 0.809 | 0.02 | NS |
| T3 (90 min) | 2 (0–4) | 2 (0–4) | 2 (0–4) | 0.271 | 0.09 | NS |
| *Within-group (Friedman)* |  |  |  |  |  |  |
| Kendall's *W* | 0.64 [0.48, 1.00] | 0.31 [0.16, 1.00] | 0.47 [0.38, 1.00] |  |  |  |
| Friedman *p* | < 0.001 | 0.027 | 0.003 |  |  |  |

KW = Kruskal–Wallis; NS = not significant; Dunn post-hoc with Holm correction.
\* Both $p = 0.025$ . <sup>†</sup> $p = 0.033$ .

Within-group analyses confirmed statistically significant pain trajectories in all groups. The tramadol group showed a gradual and highly consistent increase from T0 (median 0) to T2–T3 (median 2), with the largest concordance (Kendall’s *W* = 0.64, 95% CI [0.48, 1.00]; Friedman *χ*^2^(3) = 19.2, *p <* 0.001). The diclofenac group demonstrated a rapid step increase from T0 (median 0) to T1 (median 2), remaining stable thereafter (*W* = 0.47, 95% CI [0.38, 1.00]; *p* = 0.003). The ketorolac group exhibited a transient pain peak at T1 (median 3) followed by partial resolution to median 2 at T2–T3 (*W* = 0.31, 95% CI [0.16, 1.00]; *p* = 0.027). In all groups, final NRS values at T3 remained in the low range (median 2) (Table 2).

Given the observed age imbalance between groups (ketorolac group mean age 44.5 years vs. 32–35 years in other groups), we conducted post-hoc analyses to assess whether age influenced pain scores. Across all patients, Spearman correlation showed a moderate positive association between age and pain at T0 (*ρ* = 0.49, 95% CI [0.18, 0.72], p = 0.006), suggesting that older patients reported higher pain upon recovery room arrival. At T1, this correlation weakened and was no longer significant (*ρ* = 0.29, 95% CI [–0.08, 0.59], p = 0.12) (Figure 3).

**Figure 3:**
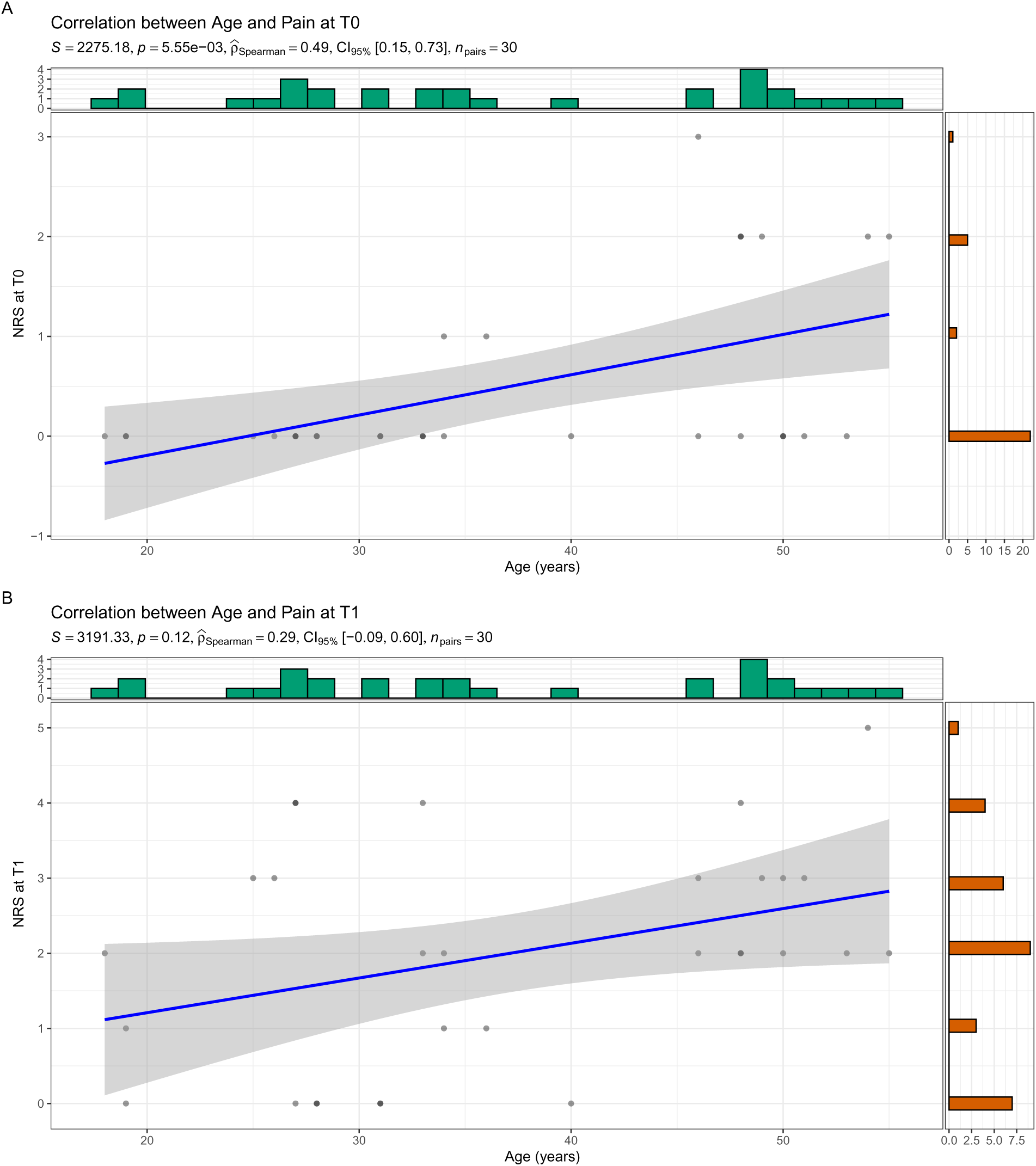
Scatter plots showing the correlation between age and NRS pain scores. At (A) T0 (recovery room arrival) and (B) T1 (30 min later). Each point represents a patient. Spearman’s *ρ* and p-values are shown. Age correlated significantly with pain at T0 (*ρ* = 0.49, p = 0.006) but not at T1 (*ρ* = 0.29, p = 0.12). Marginal histograms display variable distributions. The older age of the ketorolac group likely contributed to their higher T0 scores, but by T1 the groups converged.

To further disentangle the effects of age and treatment group, we performed linear regression with NRS as the outcome, group as a factor, and age as a covariate. At T0, after adjusting for age, the group effect was attenuated and no longer reached statistical significance (F(2,26) = 2.85, p = 0.076), while age approached significance as a predictor (*β* = 0.026, p = 0.060). At T1, neither group (F(2,26) = 2.27, p = 0.124) nor age (*β* = 0.024, p = 0.326) significantly predicted pain. These findings suggest that the early pain differences at T0 were largely attributable to the older age of the ketorolac cohort, whereas by T1 all groups converged regardless of age.

### 3.4. Verbal Rating Scale

VRS findings were consistent with the NRS data (Figure 4). At Pre, all 30 patients rated pain as “absence” across all groups, showing baseline balance. At T0, a significant between-group difference was detected (*χ*^2^(2) = 8.52, p = 0.010, Cramér’s *V* = 0.47), with 60% of ketorolac patients rating pain as “low” compared with 10% in each of the other groups. From T1 onwards, no statistically significant differences were observed (T1: p = 0.26; T2: p = 0.58; T3: p = 0.51), with the “low” category predominating in all arms (70–90% of patients by T2–T3).

**Figure 4:**
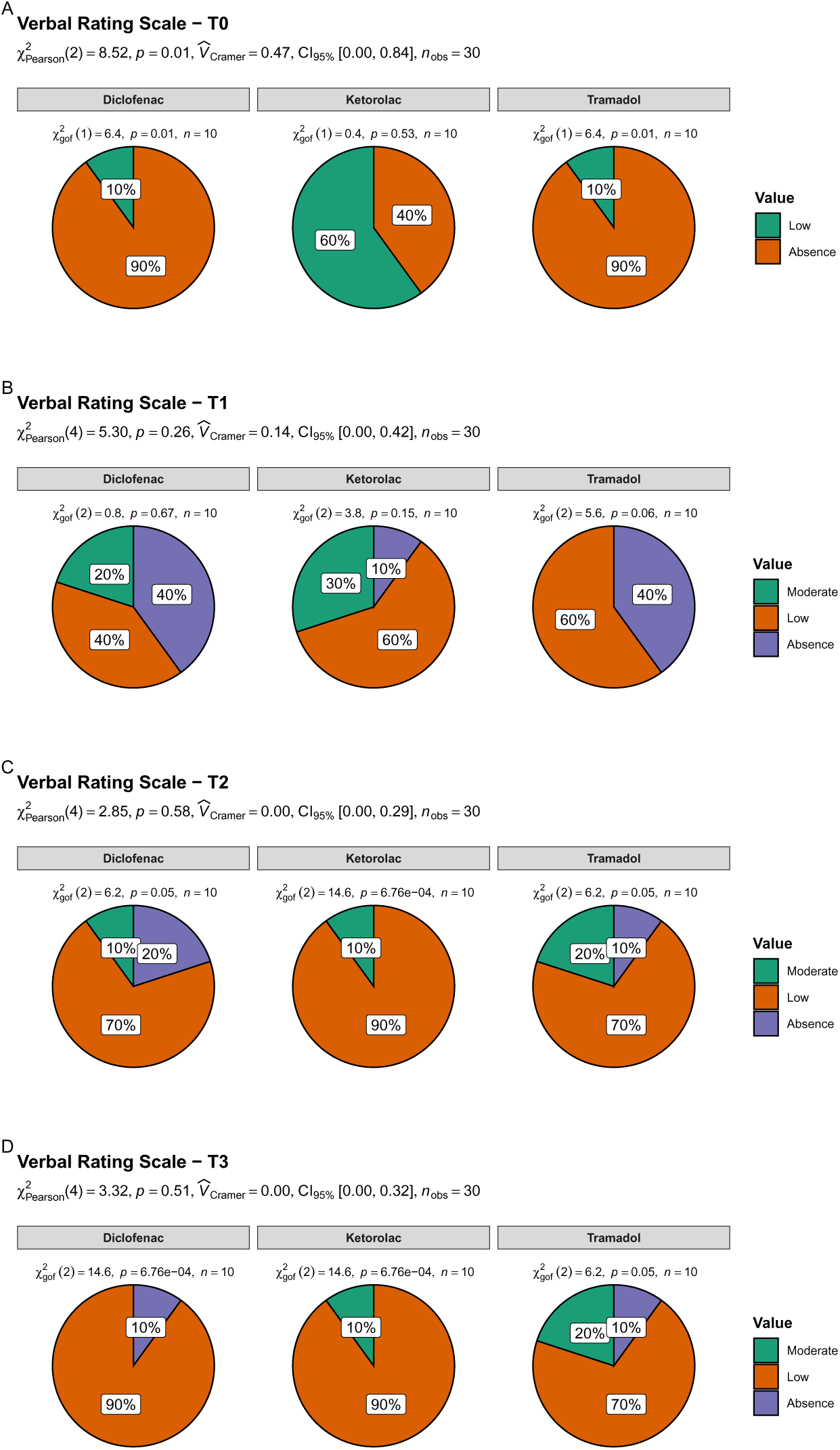
Verbal Rating Scale (VRS) Pain Categories Over Time. Stacked bar charts showing proportion of patients reporting “absence”, “low”, or “moderate” pain at each timepoint. By T2–T3, “low” pain predominates in all groups (70–90% of patients).

### 3.5. Rescue Analgesia

No rescue analgesia was required by any participant at Pre or T0. At T1, tramadol patients required no rescue doses (0/10, 0%), while 30% of the ketorolac group (3/10) and 20% of the diclofenac group (2/10) received morphine 3 mg IV; however, this between-group difference was not statistically significant (*χ*^2^(2) = 3.36, p = 0.19). At T2 and T3, rescue requirements were similarly low and comparable across groups (all p > 0.30). Total morphine consumption mirrored the number of rescue doses, with no significant between-group differences at any time point.

### 3.6. Safety

No adverse events were observed in any participant across all three groups throughout the observation period. Monitored events included hypersensitivity reactions (classified per Müller criteria), nausea, vomiting, excessive sedation, and local infusion site reactions. None occurred in any arm.

## 4. Discussion

This randomized controlled trial evaluated the analgesic equivalence of tramadol, ketorolac, and diclofenac as premedication in minimally invasive surgery. The study demonstrated that all three agents provided comparable postoperative pain control after the first 60 minutes, with no statistically significant differences in pain scores or rescue analgesic requirements at T2 (60 min) and T3 (90 min). The sample size was calculated to detect a large effect size between groups; the observed convergence of all groups to a median NRS of 2 by 60 minutes supports the hypothesis of cross-class analgesic comparability at equianalgesic doses.

The transient pain peak observed in the ketorolac group at T1 (median NRS 3) warrants interpretation. Ketorolac reaches maximum plasma concentration approximately 45–60 minutes after IV administration; therefore, pain experienced at T1 (30 minutes after recovery arrival) may partly reflect the waning of intraoperative fentanyl before ketorolac’s full analgesic effect is established. Moreover, the ketorolac cohort had a numerically older mean age (44.5 years) compared with the tramadol (32.2 years) and diclofenac (34.7 years) groups. Post-hoc analyses suggested that age correlated positively with pain at T0 (*ρ* = 0.49, p = 0.006) and that adjusting for age attenuated the group effect. Thus, the early pain peak in the ketorolac group may be partly explained by age rather than inferior efficacy. By T2–T3, all groups converged, suggesting that once established, the analgesic effect of ketorolac is similar to that of tramadol and diclofenac.

The tramadol group showed the highest within-group concordance (Kendall’s W = 0.64), suggesting a more predictable pain trajectory, whereas the ketorolac group had wider variability (W = 0.31), partially attributable to the T1 peak. The diclofenac group demonstrated an intermediate and stable profile (W = 0.47), which may reflect its dual COX-1/COX-2 inhibition providing a consistent analgesic effect once established [9, 10].

### 4.1. Comparison with Previous Studies

Our findings are broadly consistent with previous studies comparing NSAIDs and weak opioids in acute pain settings. The SPACE trial by Krebs et al. found that opioid therapy did not demonstrate superiority over NSAID therapy for chronic musculoskeletal pain [11]. Similarly, Jones et al. showed no difference in pain control between NSAIDs and opioids at one hour and at four to seven days following acute soft tissue injury [12]. While those trials addressed different pain contexts, they reinforce the view that cross-class analgesic comparability, when properly dosed, is a clinically relevant and achievable goal.

### 4.2. Strengths and Limitations

This study has several strengths. The use of a double-blind, randomised design with equianalgesic doses provides a rigorous comparison of the three agents. The sample size was calculated a priori to detect a large effect size between groups. Protocol adherence and retention were 100%, supporting the feasibility of this intervention in clinical practice.

Several limitations must be acknowledged. First, the sample size (n = 10 per group) limits statistical power for formal equivalence testing; the wide confidence intervals on effect size estimates reflect this. Second, the unintentional age imbalance between groups is a confounding variable; although post-hoc regression analyses suggested that age contributed to early pain differences, this finding is exploratory. Third, follow-up was limited to 90 minutes, which does not capture longer-term outcomes. Fourth, the study enrolled a heterogeneous minimally invasive surgical population (two different procedures), introducing variability in pain stimuli. Fifth, the absence of a placebo arm means that absolute analgesic efficacy cannot be inferred. Sixth, the trial was registered retrospectively on ClinicalTrials.gov (NCT07500454) because access to an international registry was not available at the time of study initiation. While this does not affect the integrity of the data, it is a limitation that should be considered when interpreting the results; the study nonetheless adhered to CONSORT guidelines and disclosed registration transparently.

### 4.3. Implications for Future Research

Based on the observed variance (SD = 2.1 at T2), a future non-inferiority three-arm trial with a margin of 1 NRS point, *α* = 0.05 (Bonferroni-corrected), and power = 80% would require approximately 27 patients per group (81 total); allowing for 10% attrition, 30 patients per group (90 total) is a reasonable target. Future trials should stratify randomisation by age, standardise the surgical procedure, extend follow-up to 24 hours, and consider a placebo arm or an active comparator with a validated non-inferiority margin.

## 5. Conclusion

In this randomised controlled trial of 30 patients undergoing minimally invasive surgery under balanced general anaesthesia, premedication with equianalgesic doses of tramadol 150 mg, ketorolac 60 mg or diclofenac 150 mg IV provided comparable postoperative pain control and rescue analgesic requirements over 90 minutes. All three agents were well tolerated, with no hypersensitivity reactions observed. The sample size was calculated a priori to detect a large effect size between groups, and the observed convergence of all groups to a median NRS of 2 by 60 minutes supports the hypothesis of cross-class analgesic comparability at equianalgesic doses. These findings provide clinically useful guidance for analgesic selection in minimally invasive surgery, particularly in resource-limited settings where access to potent opioids is restricted.

## 6. Authors’ Contributions

PV: conceptualization, data curation, formal analysis, investigation, methodology, project administration, resources, supervision, validation, writing-reviewing and editing; PL: data curation, formal analysis, methodology, project administration, software, supervision, validation, visualization, writing-reviewing and editing; MD: conceptualization, funding acquisition, investigation, methodology, project administration, resources, supervision, validation, writing-reviewing and editing.

All authors have read and approved the final manuscript.

## Acknowledgments

The authors acknowledge the *Instituto de Seguridad y Servicios Sociales de los Trabajadores del Estado Hospital Regional “General Ignacio Zaragoza“*, its Department of Anesthesiology, Dr. Miguel Pineda Sánchez, M.D.; Dr. Andrés Hernández Ortiz, M.D.; Dr. Kenia Olvera Guerrero, M.D.; Dr. Maria del Rosario Escalona Arroyo, M.D.; and the *Universidad Nacional Autónoma de México*.

## 9. Data Availability

Deidentified individual participant data will be made available following article publication, on GitHub <https://github.com/phabel-LD>, along with all analysis code.

